# Impact of Transjugular Intracardiac Echocardiography-Guided Self-Expandable Transcatheter Aortic Valve Implantation on Reduction of Conduction Disturbances

**DOI:** 10.1101/2023.03.28.23287887

**Authors:** Kenichi Ishizu, Shinichi Shirai, Norihisa Miyawaki, Kenji Nakano, Tadatomo Fukushima, Euihong Ko, Yasuo Tsuru, Hiroaki Tashiro, Hiroyuki Tabata, Miho Nakamura, Toru Morofuji, Takashi Morinaga, Masaomi Hayashi, Akihiro Isotani, Nobuhisa Ohno, Shinichi Kakumoto, Kenji Ando

## Abstract

**Background:** A high permanent pacemaker implantation (PPI) risk remain a concern of self-expandable transcatheter aortic valve implantation (TAVI), despite continued improvements in implantation methodology. We aimed to assess the impact of real-time direct visualization of the membranous septum using transjugular intracardiac echocardiography (ICE) during TAVI on reducing the rates of conduction disturbances including the need for PPI.

**Methods:** Consecutive patients treated with Evolut R and Evolut PRO/PRO+ from February 2017 to September 2022 were included in this study. We compared outcomes between the conventional implantation method using the 3-cusps view (“3-cusps without ICE” group), the recent method using cusp-overlap view (“cusp-overlap without ICE” group), and our novel method using ICE (“cusp-overlap with ICE” group).

**Results:** Of the 446 patients eligible for analysis, 211 (47.3%) were categorized as the “3-cusps without ICE” group, 129 (28.9%) were in the “cusp-overlap without ICE” group, and 106 (23.8%) comprised the “cusp-overlap with ICE” group. Compared with the “3-cusps without ICE” group, the “cusp-overlap without ICE” group had a smaller implantation depth (2.2 [IQR 1.0–3.5] mm vs 4.3 [IQR: 3.3–5.4], *P* <0.001) and lower 30-day PPI rates (7.0% vs 14.2%, *P* = 0.035). Compared with the “cusp-overlap without ICE” group, the “cusp-overlap with ICE” group had lower 30-day PPI rates (1.0%, *P* = 0.014), albeit with comparable implantation depths (1.9 [IQR 0.9–2.9] mm, *P* = 0.150). Multivariable analysis showed that our novel method using ICE with the cusp-overlap view was independently associated with a 30-day PPI rate reduction. There were no group differences in 30-day all-cause mortality (1.4% vs 1.6% vs 0%; *P* = 0.254).

**Conclusions:** Our novel implantation method using transjugular ICE, which enabled a real-time direct visualization of the membranous septum, achieved a predictably high position of prostheses, resulting in a substantial reduction of conduction disturbances requiring PPI after TAVI.

## Introduction

In the past decades, recommendations for transcatheter aortic valve implantation (TAVI), which was established as a therapeutic alternative to surgical aortic valve replacement (SAVR) for inoperable or high-risk patients with severe aortic stenosis (AS), have expanded to include lower-risk patients.^1–3^ However, a higher risk of conduction disturbances requiring permanent pacemaker implantation (PPI) remain a serious concern of self-expandable TAVI. Indeed, a large-scale randomized trial of low-surgical-risk patients demonstrated non-inferior outcomes of self-expandable TAVI to SAVR, but the excess of new PPI for self-expandable TAVI was noted albeit with its repositionable nature (17.4% vs 6.1%).^4^ Anatomically, the conduction system is located at the lower end of the membranous septum (MS), and high implantation is required to mitigate the risk of PPI in patients with a short MS. The recently developed cusp-overlap method has been widely used in place of the conventional 3-cusps method to achieve an accurate transcatheter heart valve (THV) implantation depth by eliminating the parallax between the aortic annulus and the delivery catheter. However, the Optimize PRO study interim analysis^5^, which examined the usefulness of the cusp-overlap method, reported a still relatively high PPI rate of 8.8%. Therefore, we utilized transjugular intracardiac echocardiography (ICE) to continuously monitor the MS during TAVI and attempted to reduce the PPI rate by repositioning the device based on the ICE findings. The aim of our study was to assess the impact of transjugular ICE-guided self-expandable TAVI on reducing the rates of post-procedural conduction disturbances, compared to those resulting from conventional implantation methods without ICE.

## Methods

### Study Population and Design

From February 2017 to September 2022, 1531 consecutive patients undergoing TAVI at the Kokura Memorial Hospital were prospectively included in an institutional database. All patients were considered as eligible candidates for TAVI rather than SAVR via consensus of the heart team, and written informed consent was obtained from all patients before the TAVI procedure. The study conformed to the principles outlined in the Declaration of Helsinki and was approved by the Institutional Review Board of Kokura Memorial Hospital. After excluding patients with balloon-expandable TAVI, previous cardiac electronic device, and previous aortic bioprosthesis, a total of 446 patients treated with contemporary repositionable self-expandable TAVI for symptomatic severe native AS were included in the final analysis (**Figure 1**).

**Figure 1.**
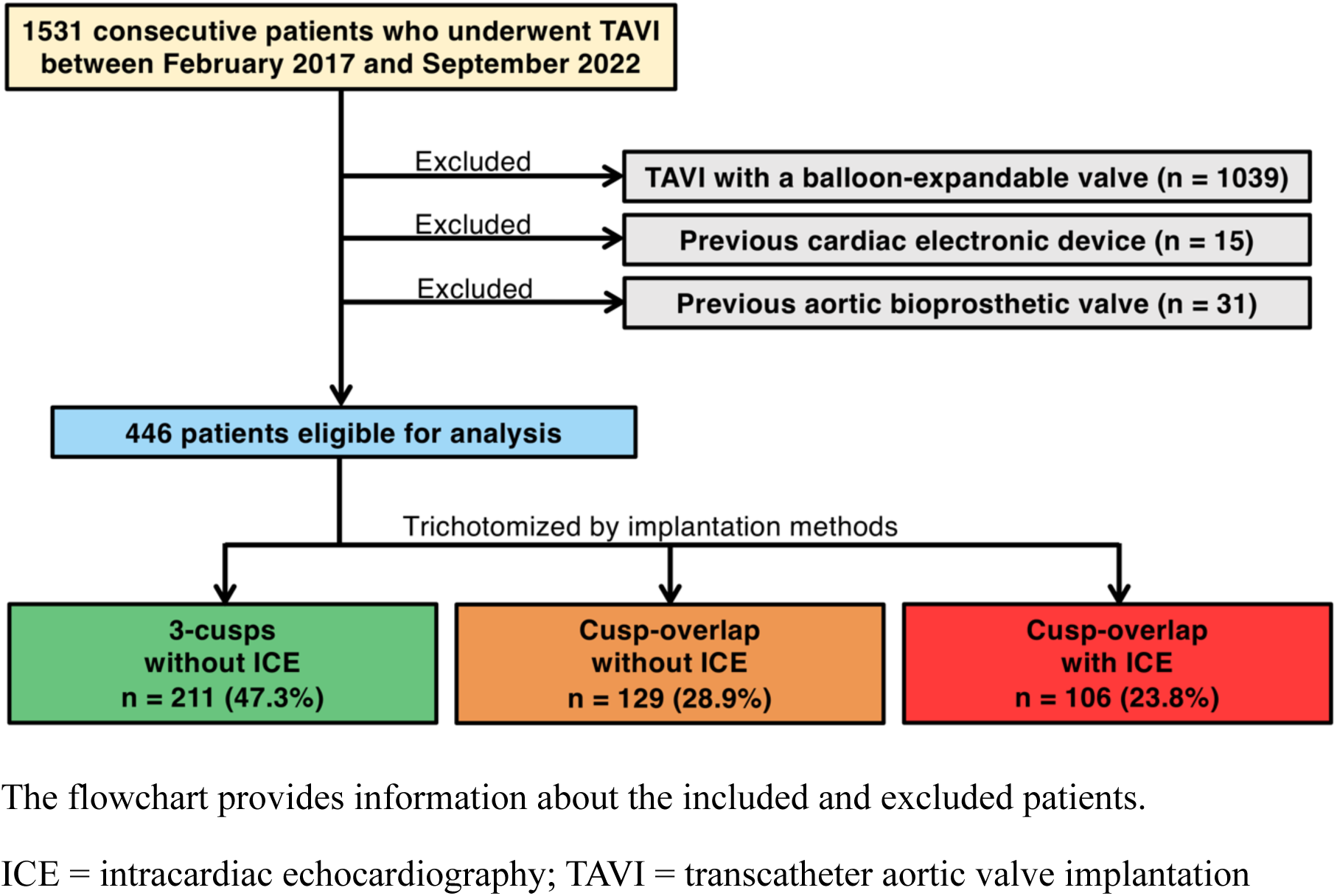
Study Flowchart The flowchart provides information about the included and excluded patients. ICE = intracardiac echocardiography; TAVI = transcatheter aortic valve implantation

### Multidetector Computed Tomographic Image Acquisition and Analysis

Computed tomographic examinations were performed as previously described.^6, 7^ All patients underwent pre-procedural electrocardiographically-gated multidetector computed tomography (MDCT) using a 256-row system (Revolution CT, GE Healthcare, Waukesha, WI) with a slice thickness of 0.625 mm and 25–70 mL of intravenously administered contrast agent (Oypalomin 350, Fuji Pharma, Tokyo, Japan). Off-line measurements were performed with 3mensio Valves software version 7.0 or 8.0 (Pie Medical Imaging, Maastricht, Netherlands). The aortic annulus and left ventricular outflow tract (LVOT) area were measured in mid-systole, and the other measurements were obtained in diastole. As previously described, the MS length was measured by determining the thinnest part of the interventricular septum on the perpendicular annular plane (axial) image, using the perpendicular crosshairs to find the corresponding stretched vessel image and using the latter to measure the perpendicular vertical distance from the annular plane to the vertex of the muscular septum (**Figure S1**).^8^

### Procedure and Implantation Technique

Evolut R and Evolut PRO/PRO+ (Medtronic, Minneapolis, MN) THVs were implanted, and the sizing of which was determined on the basis of contrast-enhanced MDCT findings. Before October 2019, 23-, 26-, and 29-mm THVs were used, and after its commercial release in October 2019, 34-mm Evolut PRO+ were also included. The femoral artery was the first-choice approach route; if femoral access was inappropriate, the iliac artery, subclavian, or direct aortic routes were considered. The 3-cusps coplanar angle was angiographically modified during the procedure based on the pre-procedural MDCT. The conventional deployment method (earlier era) includes 4 elements: 1) advancing the delivery catheter across the native valve in the 3-cusps coplanar view; 2) eliminating parallax of the delivery catheter by moving the C-arm to more left anterior oblique and/or caudal from the coplanar view; 3) deploying the THV to the point of no recapture; and 4) determining the suitability for immediate device release or device repositioning by a pre-release coaxial root angiogram. In July 2020, we introduced the systematic deployment method proposed by Tang et al.^9^, including 5 elements: 1) advancing the delivery catheter across the native valve in the cusp-overlap view, a coplanar projection achieved by overlapping the right coronary cusp and left coronary cusp; 2) deploying the THV to the point of no recapture; 3) confirming the target implantation depth from the bottom of the noncoronary cusp by a root angiogram in the cusp-overlap view; 4) assessing the implantation depth from the bottom of the left coronary cusp and the coaxiality of the delivery catheter with the aorta by a root angiogram in the 3-cusps coplanar view; and 5) determining the suitability for immediate device release or device repositioning by these two root angiograms. Moreover, in addition to the cusp-overlap technique, we have routinely used ICE during TAVI to continuously visualize the MS, the bottom of which is known to be a risky landing zone for conduction disturbances. In all cases, the AcuNav (Biosense Webster, Irvine, CA) ICE catheter was inserted from the right jugular vein, enabling real-time evaluation for not only the relationship between the THV stent frame and the MS but also the stiff wire in the left ventricle and paravalvular leakage. When the bottom of the THV was located at a deeper position than MS at the point of no recapture, we unexceptionally repositioned the THV to a higher position within the MS to prevent the stent frame from damaging the conduction system (**Figure 2**). Therefore, the patients in our study were divided into three chronological groups: 1) “3-cusps without ICE” group; 2) “cusp-overlap without ICE” group; and 3) “cusp-overlap with ICE” group (**Figure 1**).

**Figure 2.**
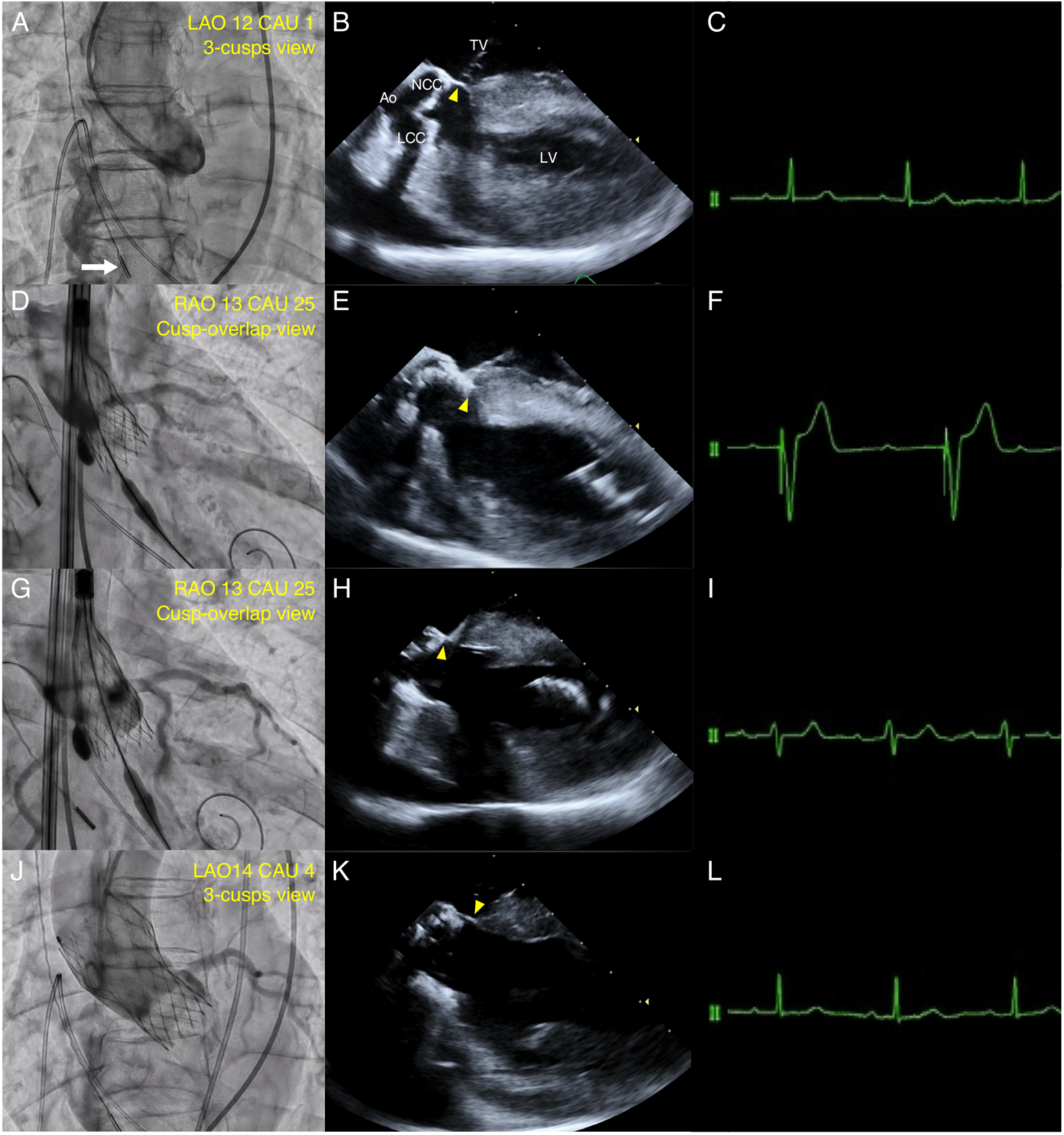
Real-Time Direct Visualization of Membranous Septum with Intracardiac Echocardiography (A) The 3-cusps coplanar angle was angiographically modified during the procedure. The white arrow indicates the position of intracardiac echocardiography (ICE) catheter. (B) Baseline ICE enabled direct visualization of membranous septum (MS) (yellow arrowhead). (C) Baseline electrocardiogram showing sinus rhythm with narrow QRS. (D) At the first attempt to deploy a 23-mm Evolut PRO+ to the point of no recapture in the cusp-overlap view, (E) ICE showed the bottom of transcatheter heart valve (THV) located at a deeper position than MS (yellow arrowhead), and (F) electrocardiogram showed complete atrioventricular block. (G) At the second attempt to the point of no recapture, (H) ICE showed the optimal THV landing in the MS (yellow arrowhead), and (I) electrocardiogram showed recovery of sinus rhythm with left bundle branch block (LBBB). (J) Final angiography showed trivial paravalvular leakage with the depth of 2 mm, and (K) ICE showed the bottom of MS remained intact (yellow arrowhead). (L) By the end of the procedure, electrocardiogram showed recovery from LBBB. Ao = aorta; LCC = left coronary cusp; LV = left ventricle; NCC = non-coronary cusp; TV = tricuspid valve.

### Electrocardiography

Following the procedure, patients had several twelve-lead electrocardiograms (ECGs) to document serial changes in conduction. Those without significant changes in cardiac conduction were routinely discharged on post-operative day five. Conduction disturbances were determined as per standard definitions by the American Heart Association, American College of Cardiology Foundation, and Heart Rhythm Society recommendations for the standardization and interpretation of ECGs.^10^ A new-onset high-grade atrioventricular block (AVB) was defined as the development of a second- or third-degree AVB on post-procedural ECGs. A new-onset left bundle branch block (LBBB) was dichotomized into persistent LBBB or transient LBBB. Persistent LBBB was defined as the development of LBBB on post-procedural ECGs that persisted to discharge.

### Outcome Measures and Follow-up

The primary outcome measure of the study was PPI within 30 days after the TAVI procedure. As specific guidelines about the indications for post-TAVI PPI are lacking,^11^ we defined the indication for PPI as the presence of persistent high-grade AVB with the agreement about the need for PPI in our electrophysiology team. We also assessed the other new-onset conduction disturbances, including LBBB, THV implantation depth, and procedural outcomes. All patients underwent up to 30 days of follow-up. Outcomes were assessed according to the Valve Academic Research Consortium (VARC)-3 criteria. ^12^

### Measurements of Prosthesis Implantation Depth

Prosthesis implantation depth was measured from an off-line evaluation of post-TAVI electrocardiographically-gated MDCT, which was routinely performed several days after the procedure in our institute to assess the positional relation of the implanted prosthesis and the surrounding structures. Off-line measurements were performed with 3mensio Valves software version 7.0 or 8.0 (Pie Medical Imaging, Maastricht, Netherlands) as with the pre-TAVI MDCT. Implantation depth was defined as the vertical distance from the native annulus to the most proximal edge to the stent frame on the same stretched vessel plane used to measure the MS length in the pre-TAVI MDCT (**Figure S2**). To evaluate intra- and inter-observer variabilities, we randomly selected a repeated measurement for a subset of 20 patients. Intra- and inter-observer agreements were almost perfect (intra-observer: intraclass correlation coefficient [ICC]: 0.96; inter-observer: ICC 0.94).

### Statistical Analysis

Categorical variables were described as number and percent and were compared using the chi-square test. Continuous variables were described as the mean ± standard deviation or median (interquartile range, IQR) and were compared using the independent Student’s *t-*test or Kruskal–Wallis test depending on their distributions. To test the predictive ability of our new implantation method using ICE for PPI, a multivariable logistic regression model was constructed, which comprised variables known to be associated with PPI based on clinical plausibility^13, 14^ or with *P* values <0.10 in the univariate analysis. Associations were expressed as odds ratios (OR) with 95% confidence interval (CI).

All statistical analyses were performed using JMP 14.2.0 (SAS Institute Inc., Cary, NC) and R version 4.0.2 (R Foundation for Statistical Computing, Vienna, Austria). A 2-tailed *P* value of 0.05 was used for significance testing.

## Results

### Baseline Patient Characteristics

Of the 446 patients eligible for analysis, 211 (47.3%) were categorized as the “3-cusps without ICE” group, 129 (28.9%) were included in the “cusp-overlap without ICE” group, and 106 (23.8%) comprised the “cusp-overlap with ICE” group. The main baseline clinical characteristics are presented in **Table 1**. The mean age of our study population was 85 (IQR: 82–89) years, 24% of patients were male, and the median Society of Thoracic Surgeons (STS) risk score was 5.3 (IQR: 3.7–8.0). Patient demographics were comparable between the three groups, except for the prevalence of male (19.0% vs 30.2% vs 27.4%; *P* = 0.042) and the Society of Thoracic Surgeons (STS) risk score (6.1 [IQR: 4.1–9.4] % vs 4.7 [IQR: 3.5–7.0] % vs 5.0 [IQR: 3.7–7.1] %; *P* = 0.001). The MS length was similar between the three groups (*P* = 0.824).

**Table 1.** Baseline Characteristics

|  | Implantation methods |  |  | *P*<br>value |
| --- | --- | --- | --- | --- |
|  | 3-cusps<br>without ICE<br>(N = 211) | Cusp-overlap<br>without ICE<br>(N = 129) | Cusp-overlap<br>with ICE<br>(N = 106) |  |
| Demographics |  |  |  |  |
| Age, years | 86 (82–89) | 85 (81–88) | 85 (81–89) | 0.540 |
| Male | 40 (19.0) | 39 (30.2) | 29 (27.4) | 0.042 |
| BMI, kg/m<sup>2</sup> | 21.4 (19.6–23.7) | 22.1 (19.9–24.8) | 21.2 (18.9–24.4) | 0.187 |
| Clinical Frailty Scale ≥4 | 90 (42.7) | 53 (41.1) | 52 (49.1) | 0.431 |
| NYHA functional class III/IV | 72 (34.1) | 35 (27.1) | 32 (30.2) | 0.387 |
| STS risk score, % | 6.1 (4.1–9.4) | 4.7 (3.5–7.0) | 5.0 (3.7–7.1) | 0.001 |
| Comorbidities |  |  |  |  |
| Hypertension | 174 (86.5) | 102 (79.1) | 87 (82.1) | 0.726 |
| Dyslipidemia | 102 (48.3) | 69 (53.5) | 52 (49.1) | 0.638 |
| Diabetes | 39 (18.5) | 33 (25.6) | 28 (26.4) | 0.164 |
| Atrial fibrillation | 40 (19.0) | 31 (24.0) | 20 (18.9) | 0.488 |
| Coronary artery disease | 46 (21.8) | 65 (50.4) | 40 (37.7) | <0.001 |
| Previous coronary bypass | 4 (1.9) | 3 (2.3) | 1 (0.9) | 0.695 |
| Peripheral artery disease | 13 (6.2) | 11 (8.5) | 11 (10.4) | 0.401 |
| Chronic kidney disease | 133 (63.0) | 87 (67.4) | 73 (68.9) | 0.519 |
| Chronic pulmonary disease | 21 (10.0) | 13 (10.2) | 11 (10.4) | 0.993 |
| Cerebrovascular disease | 20 (89.5) | 10 (7.8) | 9 (8.5) | 0.855 |
| Active cancer | 10 (4.7) | 8 (6.2) | 4 (3.8) | 0.685 |
| **Electrocardiographic data** |  |  |  |  |
| Persistent AF | 20 (9.5) | 16 (12.4) | 12 (11.3) | 0.685 |
| PQ interval, ms * | 176 (158–198) | 174 (155–194) | 173 (158–196) | 0.920 |
| First-degree AVB * | 44 (23.0) | 22 (19.5) | 20 (21.3) | 0.761 |
| QRS duration, ms | 92 (86–102) | 92 (84–100) | 91 (86–105) | 0.564 |
| LBBB | 9 (4.3) | 2 (1.6) | 3 (2.8) | 0.343 |
| RBBB | 23 (10.9) | 9 (7.0) | 14 (13.2) | 0.259 |
| **Echocardiographic data** |  |  |  |  |
| AVA, cm<sup>2</sup> | 0.62 (0.50–0.71) | 0.63 (0.52–0.73) | 0.60 (0.49–0.74) | 0.091 |
| Indexed AVA, cm<sup>2</sup>/m<sup>2</sup> | 0.47 (0.40–0.50) | 0.44 (0.38–0.53) | 0.44 (0.32–0.52) | 0.448 |
| Mean aortic gradient, mmHg | 54.2 (41.0–69.9) | 51.1 (38.8–64.7) | 53.2 (41.5–70.3) | 0.083 |
| LVEF, % | 63.4 (59.9–66.0) | 63.1 (57.6–65.5) | 63.0 (56.7–66.4) | 0.319 |
| AR ≥ moderate | 6 (2.8) | 4 (3.1) | 9 (8.5) | 0.071 |
| MR ≥ moderate | 10 (4.7) | 8 (6.2) | 9 (8.5) | 0.431 |
| TR ≥ moderate | 4 (1.9) | 6 (4.7) | 8 (7.6) | 0.050 |
| SPAP, mmHg | 31.0 (26.0–37.5) | 31.0 (28.0–39.0) | 32.0 (26.6–38.1) | 0.436 |
| **MDCT data** |  |  |  |  |
| Annulus area, mm<sup>2</sup> | 375.6 (340.2–425.0) | 386.9 (341.8–442.0) | 374.0 (329.2–427.2) | 0.094 |
| Annulus perimeter, mm | 69.6 (66.1–74.6) | 70.6 (67.3–75.9) | 70.7 (66.0–76.0) | 0.128 |
| LVOT area, mm<sup>2</sup> | 353.3 (301.9–426.4) | 369.2 (319.6–428.3) | 356.0 (300.7–440.7) | 0.117 |
| STJ height, mm | 18.0 (16.1–19.6) | 18.1 (16.5–20.1) | 17.9 (16.3–20.0) | 0.717 |
| STJ diameter, mm | 24.1 (22.5–26.0) | 24.3 (22.7–27.0) | 24.3 (22.9–26.2) | 0.293 |
| Mean SOV diameter, mm | 28.5 (27.1–30.2) | 29.2 (27.2–31.3) | 28.5 (27.5–30.9) | 0.144 |
| LCA height, mm | 13.3 (11.9–14.6) | 13.4 (11.9–14.9) | 13.6 (12.1–14.7) | 0.222 |
| RCA height, mm | 14.5 (12.6–16.4) | 14.9 (12.8–17.3) | 13.9 (12.5–16.1) | 0.152 |
| LVOT calcification ≥ moderate | 21 (10.0) | 15 (11.6) | 10 (9.4) | 0.838 |
| MS length, mm | 2.5 (1.7–3.6) | 2.7 (1.9–3.5) | 2.5 (1.8–3.2) | 0.824 |

Values are n (%) or median (interquartile range). \* Patients with persistent atrial fibrillation were excluded. AF = atrial fibrillation; AR = aortic regurgitation; AVA = aortic valve area; BMI = body mass index; LBBB = left bundle branch block; LCA = left coronary artery; ICE = intracardiac echocardiography; LVEF = left ventricular ejection fraction; LVOT = left ventricular outflow tract; MDCT = multidetector computed tomography; MR = mitral regurgitation; MS = membranous septum; NYHA = New York Heart Association; RBBB = right bundle branch block; RCA = right coronary artery; SOV = sinus of Valsalva; SPAP = systolic pulmonary artery pressure; STJ = sinotubular junction; STS = Society of Thoracic Surgeons; TR = tricuspid regurgitation.

### Procedural Characteristics

Procedural characteristics are listed in **Table 2**. Almost all (92.2%) patients underwent TAVI via the transfemoral approach. Overall, 59.9% received a 26-mm THV, 22.0% received a 29-mm THV, and 16.8% received a 23-mm THV. The degree of oversizing and the rate of pre- and post-dilatation did not differ between the three groups. General anesthesia was more frequently performed in the “3-cusps without ICE” group, compared with the other two groups. The procedure time and contrast volume were also greater in the “3-cusps without ICE” group.

**Table 2.** Procedural Characteristics

|  | Implantation methods |  |  | *P*<br>value |
| --- | --- | --- | --- | --- |
|  | 3-cusps<br>without ICE<br>(N = 211) | Cusp-overlap<br>without ICE<br>(N = 129) | Cusp-overlap<br>with ICE<br>(N = 106) |  |
| Local anesthesia | 151 (71.6) | 113 (87.6) | 99 (93.4) | <0.001 |
| **Access site** |  |  |  |  |
| Transfemoral | 188 (89.1) | 123 (95.4) | 100 (94.3) | 0.070 |
| Alternative | 23 (10.9) | 6 (4.7) | 6 (5.7) |  |
| **Prosthesis size** |  |  |  |  |
| 23 mm | 35 (16.6) | 21 (16.3) | 19 (17.9) | 0.167 |
| 26 mm | 124 (58.8) | 80 (62.0) | 63 (59.4) |  |
| 29 mm | 52 (24.6) | 26 (20.2) | 20 (18.9) |  |
| 34 mm | 0 (0) | 2 (1.6) | 4 (3.8) |  |
| Oversizing by perimeter, % | 17.4 (13.0–20.3) | 16.8 (12.8–20.7) | 16.5 (12.5–20.6) | 0.283 |
| Pre-dilatation | 206 (97.6) | 128 (99.2) | 105 (99.1) | 0.434 |
| Post-dilatation | 41 (19.4) | 31 (24.0) | 20 (18.9) | 0.523 |
| Procedure time, min | 60 (51–76) | 49 (43–61) | 49 (42–60) | <0.001 |
| Contrast volume, ml | 141 (110–174) | 80 (68–93) | 73 (60–88) | <0.001 |

Values are n (%) or median (interquartile range).
ICE = intracardiac echocardiography.

### Outcomes

Procedural clinical outcomes are shown in **Table 3**. Implantation depth was smaller in the “cusp-overlap without ICE” group compared with the “3-cusps without ICE” group (2.2 [IQR: 1.0–3.5] mm vs 4.3 [IQR: 3.3–5.4] mm; *P* <0.001). Moreover, the difference between the MS length and implantation depth (ΔMSID) was larger in the “cusp-overlap without ICE” group compared with the “3-cusps without ICE” group (0.4 [IQR: −0.4–1.6] mm vs −1.2 [IQR: −3.2–0.3] mm; *P* <0.001). Although a comparable median ΔMSID was observed between the “cusp-overlap without ICE” group and the “cusp-overlap with ICE” group, the variation in the ΔMSID was obviously smaller in the “cusp-overlap with ICE” group (**Figure 3**). Technical success and device success were achieved respectively in 437 (98.0%) and 414 (92.8%) patients, with significantly higher success rates across the three groups. The rates of peri-procedural complications were low and comparable between the three groups without Type 3 (life-threatening) or 4 (leading to death) bleeding (3.8% vs 2.3% vs 0%; *P* = 0.036). All-cause 30-day mortality also did not differ between the three groups (*P* = 0.254).

**Table 3.**
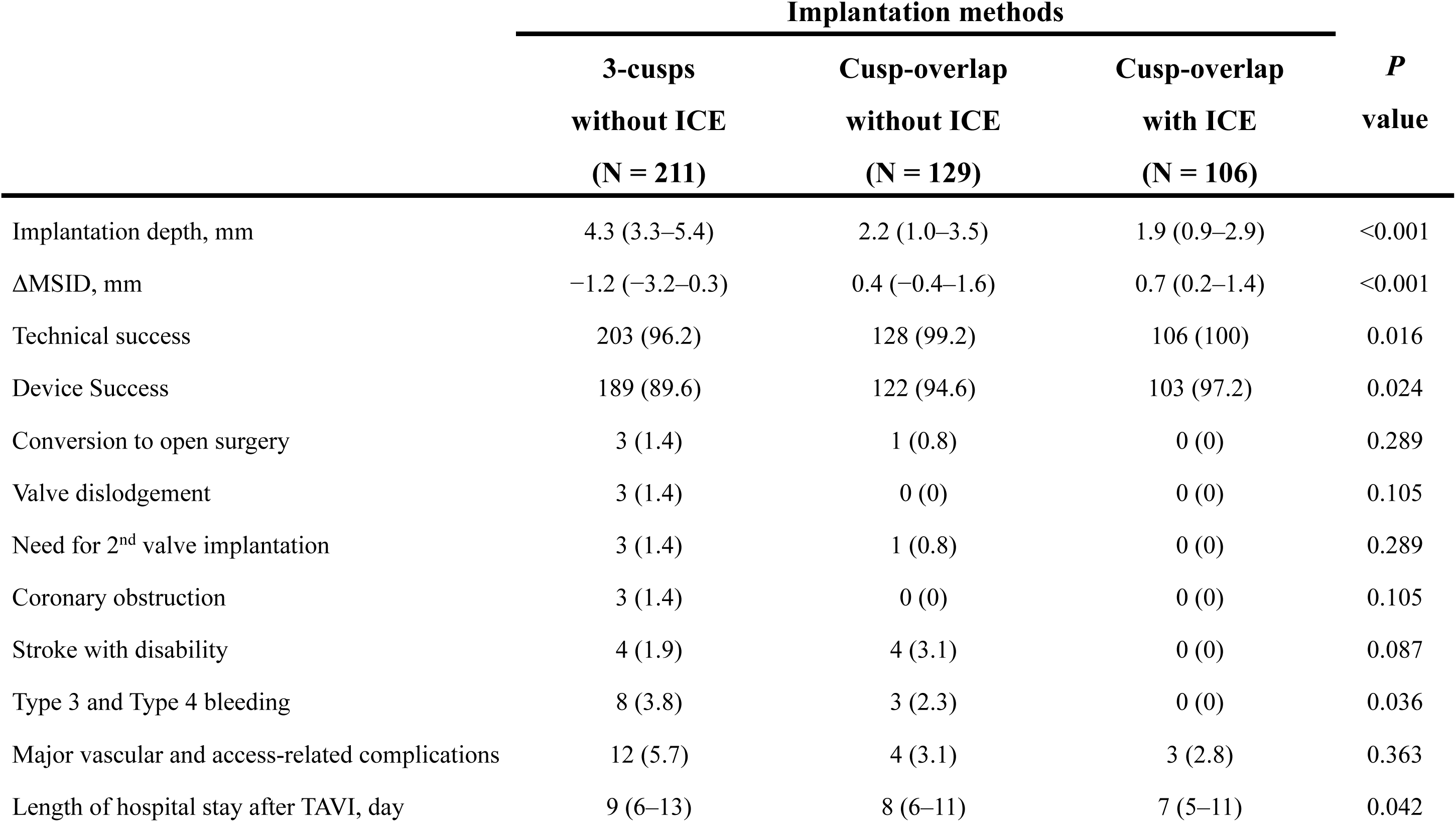
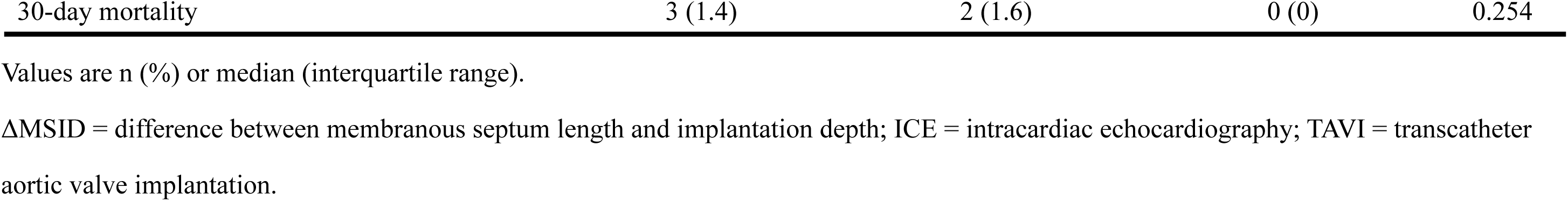
Clinical Outcomes

|  | Implantation methods |  |  | *P*<br>value |
| --- | --- | --- | --- | --- |
|  | 3-cusps | Cusp-overlap | Cusp-overlap |  |
|  | without ICE<br>(N = 211) | without ICE<br>(N = 129) | with ICE<br>(N = 106) |  |
| Implantation depth, mm | 4.3 (3.3–5.4) | 2.2 (1.0–3.5) | 1.9 (0.9–2.9) | <0.001 |
| ΔMSID, mm | –1.2 (–3.2–0.3) | 0.4 (–0.4–1.6) | 0.7 (0.2–1.4) | <0.001 |
| Technical success | 203 (96.2) | 128 (99.2) | 106 (100) | 0.016 |
| Device Success | 189 (89.6) | 122 (94.6) | 103 (97.2) | 0.024 |
| Conversion to open surgery | 3 (1.4) | 1 (0.8) | 0 (0) | 0.289 |
| Valve dislodgement | 3 (1.4) | 0 (0) | 0 (0) | 0.105 |
| Need for 2<sup>nd</sup> valve implantation | 3 (1.4) | 1 (0.8) | 0 (0) | 0.289 |
| Coronary obstruction | 3 (1.4) | 0 (0) | 0 (0) | 0.105 |
| Stroke with disability | 4 (1.9) | 4 (3.1) | 0 (0) | 0.087 |
| Type 3 and Type 4 bleeding | 8 (3.8) | 3 (2.3) | 0 (0) | 0.036 |
| Major vascular and access-related complications | 12 (5.7) | 4 (3.1) | 3 (2.8) | 0.363 |
| Length of hospital stay after TAVI, day | 9 (6–13) | 8 (6–11) | 7 (5–11) | 0.042 |
| 30-day mortality | 3 (1.4) | 2 (1.6) | 0 (0) | 0.254 |

Values are n (%) or median (interquartile range). ΔMSID = difference between membranous septum length and implantation depth; ICE = intracardiac echocardiography; TAVI = transcatheter aortic valve implantation.

**Figure 3.**
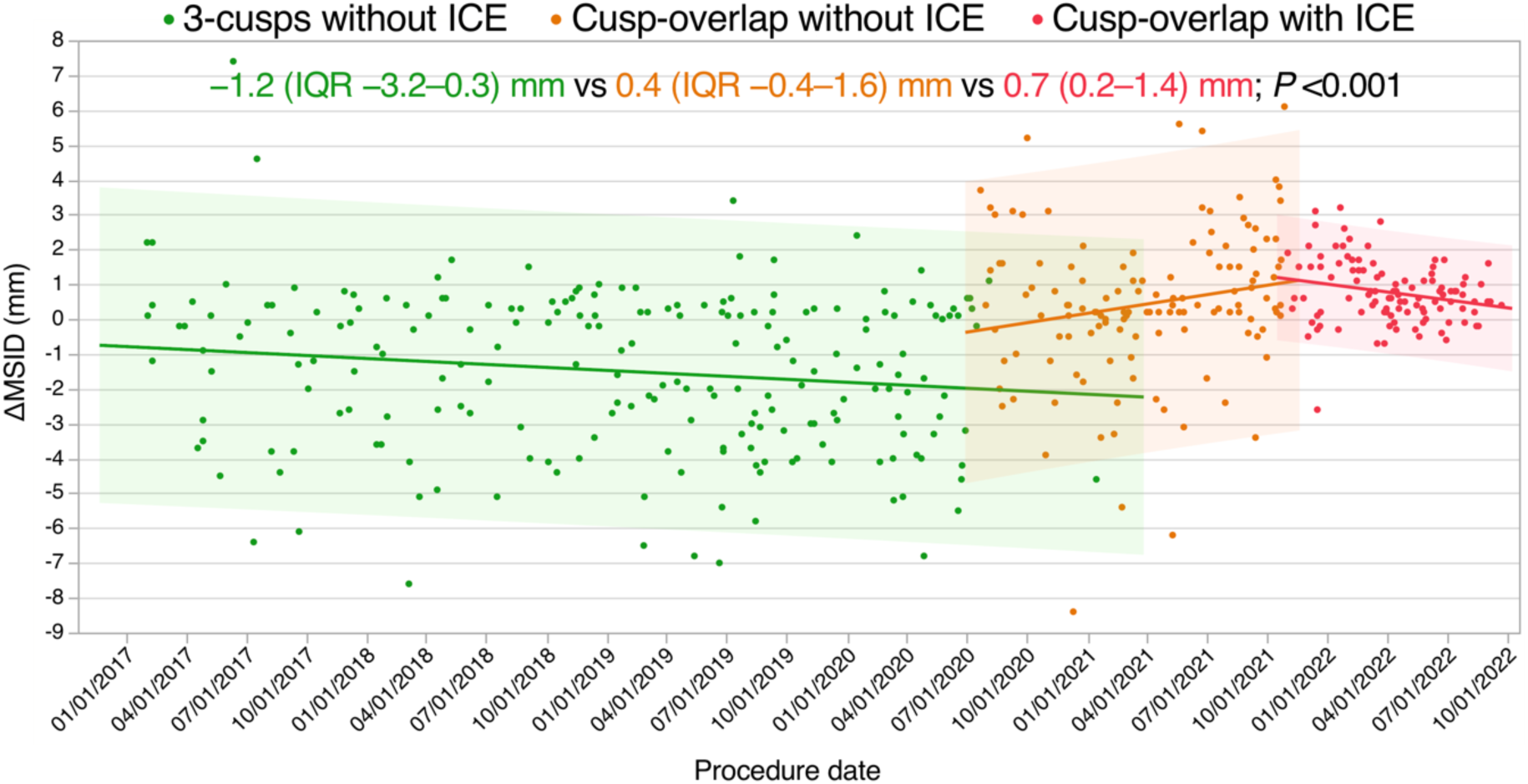
Temporal Trends of the Difference Between Membranous Septum Length and Implantation Depth (ΔMSID) with Each Technique. Filled circles indicate the final implantation depths in millimeters according to the timing of procedure. Bold lines represent the regression lines of temporal trends of median implantation depth. ICE = intracardiac echocardiography.

New-onset conduction disturbances after the TAVI procedure are detailed in **Table 4**. Within 30 days, a total of 40 patients (9.0%) underwent PPI 5 (IQR: 3–6) days after TAVI. The indication for PPI was a high-degree AVB in 3 patients and complete AVB in 37 patients. As a whole, the rate of a 30-day PPI was substantially reduced with the temporal change in implantation methods (“3-cusps without ICE” group vs “cusp-overlap without ICE” group vs “cusp-overlap with ICE” group: 14.2% vs 7.0% vs 0.9%, respectively; *P* <0.001). In addition, significant differences were observed between adjacent groups (“3-cusps without ICE” group vs “cusp-overlap without ICE” group: *P* = 0.035; “cusp-overlap without ICE” group vs “cusp-overlap with ICE” group: *P* = 0.014) (**Figure 4A**). In the “cusp-overlap with ICE” group, a THV bottom landing in the MS was achieved in all patients except for only one who underwent a deep dive of the THV into the left ventricle and consequently required a PPI 4 days after TAVI. The incidence of new-onset persistent LBBB at discharge was also lower across the groups (21.8% vs 12.4% vs 3.8%; *P* <0.001). In the multivariable analysis, compared with the “cusp-overlap without ICE” method, the “cusp-overlap with ICE” method was independently associated with a lower 30-day PPI requirement (adjusted OR, 0.07 [95% CI, 0.01–0.64]; *P* = 0.018). Baseline right bundle branch block (RBBB) (adjusted OR, 8.61 [95% CI, 3.49–21.24]; *P* <0.001) and short MS length (adjusted OR per 1 mm decrease, 1.82 [95% CI, 1.30–2.54]; *P* <0.001) also predicted a higher risk of 30-day PPI (**Table 5**). As post hoc analyses, we assessed the impact of each implantation method on 30-day PPI in the subgroups at high risk for conduction disturbances. The results showed a significantly lower 30-day PPI rate in the “cusp-overlap with ICE” group compared with the “cusp-overlap without ICE” group also in patients with baseline RBBB (7.1% vs 44.4%; *P* = 0.034) (**Figure 4B**) or with an MS length < 2.5 mm (1.9% vs 12.7%; *P* = 0.024) (**Figure 4C**).

**Table 4.** New-Onset Conduction Abnormalities and Indications for Pacemaker Implantation

|  | Implantation methods |  |  | *P*<br>value |
| --- | --- | --- | --- | --- |
|  | 3-cusps | Cusp-overlap | Cusp-overlap |  |
|  | without ICE<br>(N = 211) | without ICE<br>(N = 129) | with ICE<br>(N = 106) |  |
| 30-day PPI | 30 (14.2) | 9 (7.0) | 1 (0.9) | <0.001 |
| For advanced second-degree AVB | 2 (0.9) | 1 (0.8) | 0 (0) | 0.613 |
| For complete AVB | 28 (13.3) | 8 (6.2) | 1 (0.9) | <0.001 |
| New-onset LBBB | 83 (39.3) | 32 (24.8) | 26 (24.5) | 0.004 |
| Transient | 37 (17.5) | 16 (12.4) | 22 (20.8) | 0.209 |
| Persistent | 46 (21.8) | 16 (12.4) | 4 (3.8) | <0.001 |

Values are n (%).
AVB = atrioventricular block; ICE = intracardiac echocardiography; LBBB = left bundle branch block; PPI = permanent pacemaker implantation.

**Table 5.** Predictors of Permanent Pacemaker Requirement at 30 days

|  | Univariate Analysis |  |  | Multivariable Analysis |  |  |
| --- | --- | --- | --- | --- | --- | --- |
|  | HR | 95% CI | *P* value | HR | 95% CI | *P* value |
| **Implantation methods** |  |  |  |  |  |  |
| 3-cusps without ICE | 2.21 | 1.01–4.82 | 0.046 | 1.53 | 0.65–3.64 | 0.332 |
| Cusp-overlap without ICE | ... | ... | Reference | ... | ... | Reference |
| Cusp-overlap with ICE | 0.13 | 0.02–0.79 | 0.025 | 0.07 | 0.01–0.64 | 0.018 |
| **Adjusting factors** |  |  |  |  |  |  |
| Age (per 10 years increase) | 1.30 | 0.70–2.42 | 0.402 | ... | ... | ... |
| Male gender | 0.64 | 0.27–1.49 | 0.283 | ... | ... | ... |
| BMI (per 1 kg/m<sup>2</sup> increase) | 0.97 | 0.89–1.07 | 0.567 | ... | ... | ... |
| STS score ≥ 8% | 1.72 | 0.86–3.42 | 0.125 | ... | ... | ... |
| Baseline RBBB | 7.26 | 3.47–15.17 | <0.001 | 8.61 | 3.49–21.24 | <0.001 |
| Baseline first-degree AVB | 1.86 | 0.89–3.88 | 0.097 | 1.77 | 0.75–4.17 | 0.190 |
| Oversizing by perimeter (per 10% increase) | 1.03 | 0.89–1.19 | 0.725 | ... | ... | ... |
| LVOT area (per 10 mm<sup>2</sup> decrease) | 1.03 | 0.99–1.07 | 0.085 | 1.02 | 0.98–1.06 | 0.273 |
| MS length (per 1 mm decrease) | 1.98 | 1.44–2.71 | <0.001 | 1.82 | 1.30–2.54 | <0.001 |
| Pre-dilatation | 0.54 | 0.15–1.92 | 0.372 | ... | ... | ... |
| Post-dilatation | 2.49 | 0.90–6.88 | 0.101 | ... | ... | ... |

AVB = atrioventricular block; CI = confidence interval; BMI = body mass index; HR = hazard ratio; ICE = intracardiac echocardiography; LVOT = left ventricular outflow tract; MS = membranous septum; RBBB = right bundle branch block; STS = Society of Thoracic Surgery.

**Figure 4.**
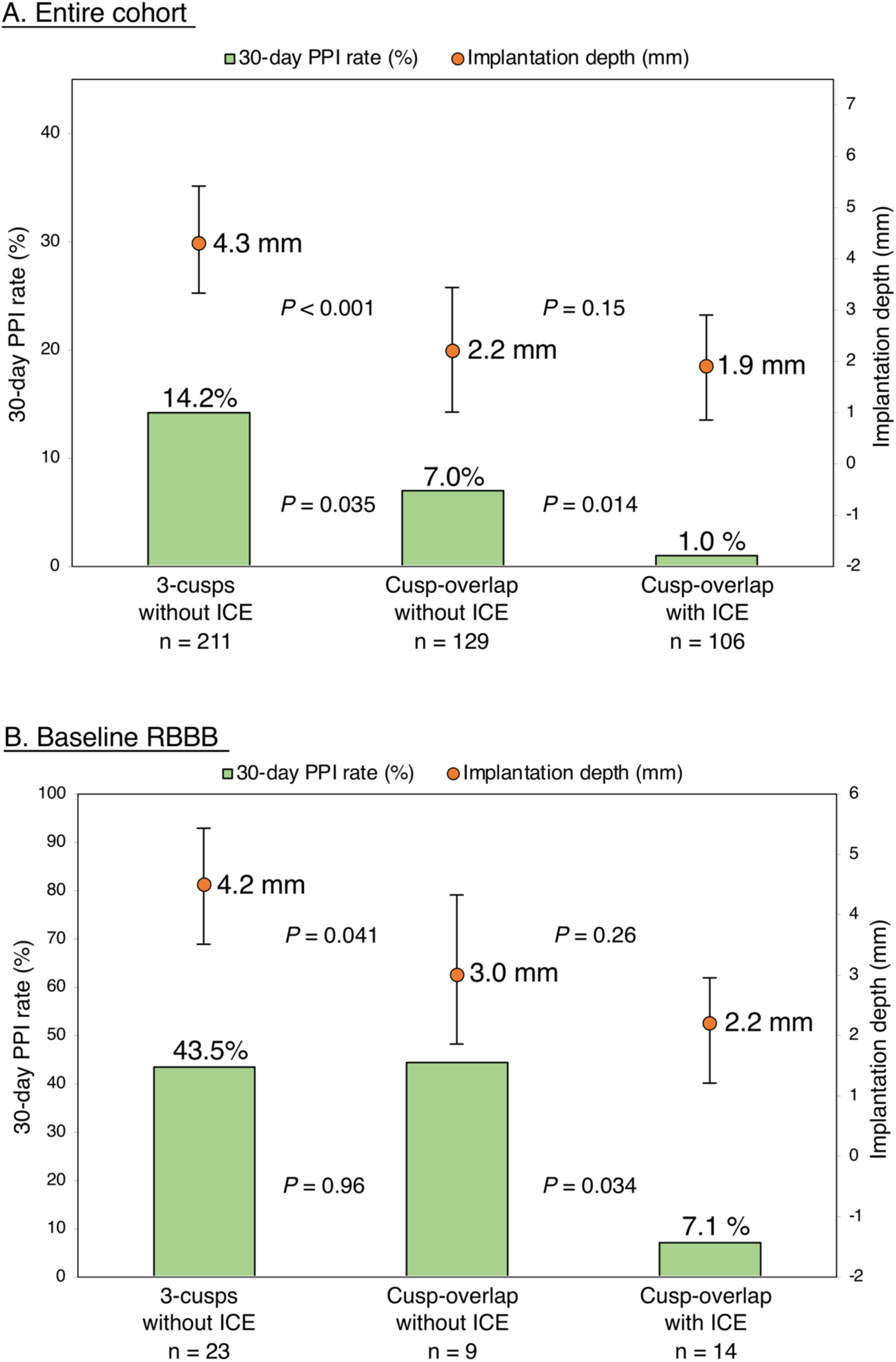
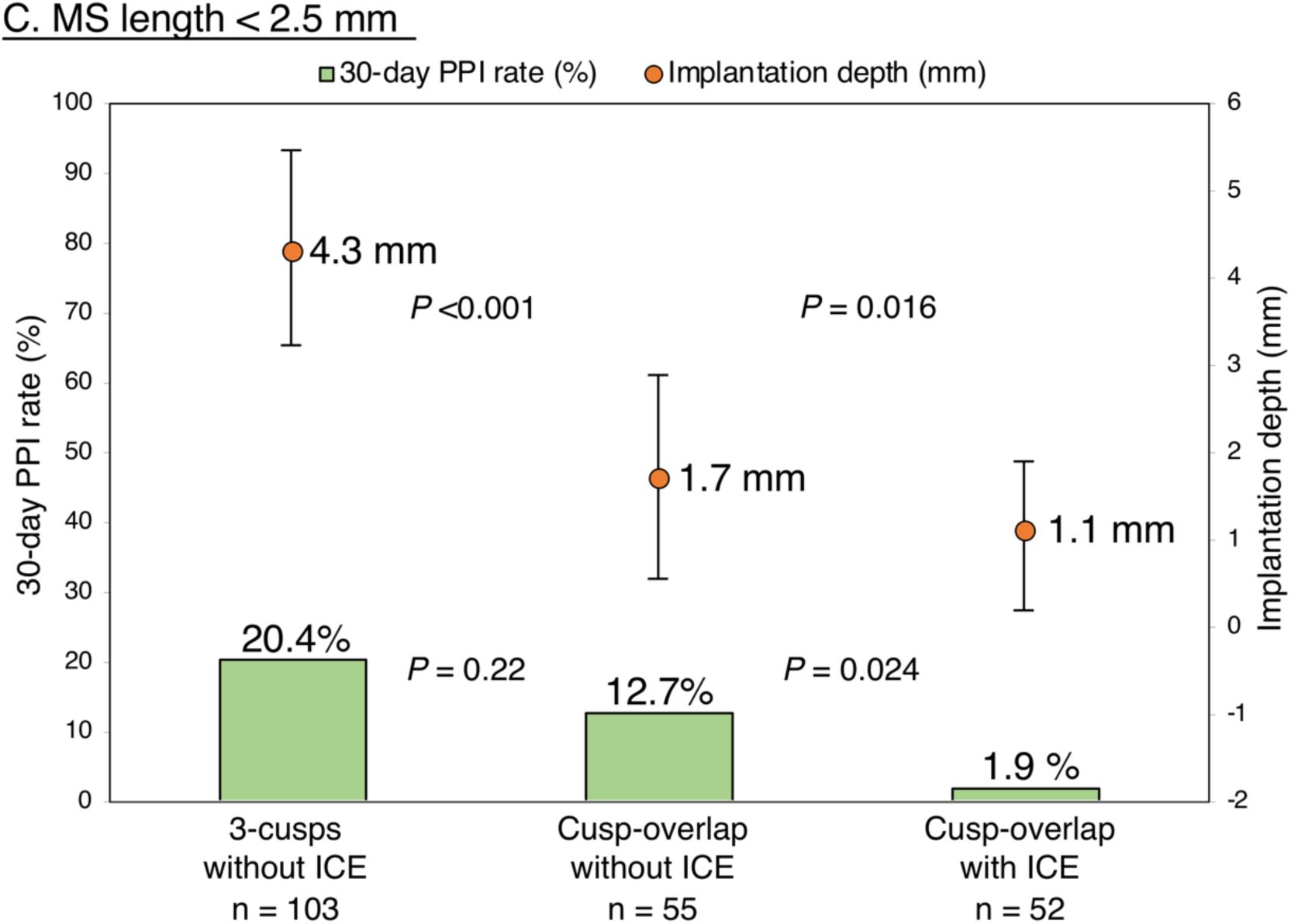
Implantation Depths and New-Onset Permanent Pacemaker Implantation (PPI) Rates for Each Implantation Method. The orange dots show implantation depth. The green columns show the rates of new-onset 30-day PPI in the entire cohort (A), in those with right bundle branch block (RBBB) (B), and in those with membranous septum (MS) length < 2.5 mm (C). The error bars indicate the interquartile range. ICE = intracardiac echocardiography

With regard to 30-day echocardiographic outcomes (**Table 6**), the “3-cusps without ICE” group demonstrated a lower indexed effective orifice area (EOA) (1.77 [IQR: 1.50– 2.03] cm^2^ vs 1.97 (IQR: 1.68–2.26) cm^2^ vs 1.93 [IQR: 1.63–2.22] cm^2^; *P* <0.001) and a higher incidence of patient-prosthesis mismatch (PPM) (5.7% vs 1.6% vs 0.9%; *P* = 0.027). In contrast, there were no differences in the rates of mild AR (56.9% vs 52.7% vs 54.7%; *P* = 0.703) or moderate to severe AR (0.5% vs 0.8% vs 0.9%; *P* = 0.878).

**Table 6.** Echocardiographic Outcomes

|  | Implantation methods |  |  | *P*<br>value |
| --- | --- | --- | --- | --- |
|  | 3-cusps | Cusp-overlap | Cusp-overlap |  |
|  | without ICE<br>(N = 211) | without ICE<br>(N = 129) | with ICE<br>(N = 106) |  |
| EOA, cm<sup>2</sup> | 1.77 (1.50-2.03) | 1.97 (1.68-2.26) | 1.93 (1.63-2.22) | <0.001 |
| Indexed EOA, cm<sup>2</sup> | 1.29 (1.06-1.48) | 1.34 (1.18-1.56) | 1.33 (1.16-1.55) | 0.013 |
| PPM | 12 (5.7) | 2 (1.6) | 1 (0.9) | 0.027 |
| Moderate PPM | 11 (5.2) | 2 (1.6) | 1 (0.9) | 0.047 |
| Severe PPM | 1 (0.5) | 0 | 0 | 0.473 |
| Mean gradient, mmHg | 10.3 (8.1-12.7) | 9.3 (6.6-11.3) | 9.3 (7.4-12.4) | 0.015 |
| LVEF, % | 63.0 (59.6-65.7) | 62.9 (58.6-65.1) | 63.2 (57.1-66.5) | 0.482 |
| Mild AR | 120 (56.9) | 68 (52.7) | 58 (54.7) | 0.703 |
| Moderate to severe AR | 1 (0.5) | 1 (0.8) | 1 (0.9) | 0.878 |

Values are n (%) or median (interquartile range).
AR = aortic regurgitation; EOA = effective orifice area; ICE = intracardiac echocardiography; LVEF = left ventricular ejection fraction; PPM =
prosthesis-patient mismatch.

## Discussion

In this study, we analyzed the impact of the novel implantation method using transjugular ICE on reducing the occurrence of conduction disturbances and PPI after the repositionable self-expandable TAVI. The main findings of our study are as follows: 1) Compared with the 3-cusps view only, the combined use of cusp-overlap view led to a higher THV position and a lower rate of conduction disturbances; 2) Most importantly, guidance with a transjugular ICE, which enabled a real-time visualization of MS, provided a significant reduction in conduction disturbances, albeit with a comparable THV implantation depth even in patients who benefited from the cusp-overlap method; and 3) Multivariable analysis also showed our novel technique, adding transjugular ICE to the cusp-overlap method, was independently associated with a 30-day PPI rate reduction following TAVI.

Self-expandable TAVI generally carries high risks for post-procedural PPI (17.4%) and new-onset LBBB (25.8%).^4, 8^ As TAVI indications are expanding to include younger patients with a low surgical risk, these conduction disturbances relevant to long-term adverse outcomes have to be promptly addressed.^15, 16^ The procedural steps of balloon-expandable THVs have remained identical for several years, whereas the deployment methods for self-expandable THV platforms continue to improve. Recently, two important modifications to the self-expandable TAVI procedure have been proposed. The first is the cusp-overlap technique advocated by Tang et al. in 2018, in which the C-arm is positioned to superimpose the left coronary cusp over the right coronary cusp and isolate the non-coronary cusp. The conventional 3-cusps view, although popular for the implantation of THVs, provides a high degree of parallax between the aortic annulus and delivery catheter with a markedly foreshortened LVOT, resulting in an inaccurate deeper position of the THV. In contrast, the cusp-overlap view avoids the parallax of the delivery catheter and foreshortening of the LVOT, and helps the operators achieve accurate THV deployment. Indeed, in our study, compared with the 3-cusps view only, the combined use of cusp-overlap view allowed a higher THV position with a lower rate of new-onset 30-day PPI (7.0% vs 14.2%). This result dovetails with those from the Optimize PRO study interim analysis^5^; however, the persistently high PPI rate as compared with that carried by SAVR remains a concern.^17^ The second is the pre-procedural assessment of the MS length for a risk stratification of conduction disturbances following TAVI. The use of MS length as an anatomic surrogate of the distance between the aortic annulus and the bundle of His was first reported by Hamdan et al. in 2015,^18^ and many studies since then have supported its importance.^8, 19, 20^ In particular, Jilaihawi et al. demonstrated that the approach aiming for implantation depth in relation to the non-coronary cusp less than MS length achieved very low and predictable PPI rates (3.0%) after self-expandable TAVI. However, in fact, the MS length may approach the minimum resolution of CT, particularly in cases with shorter MS lengths, which can potentially lead to measurement errors. More importantly, fluoroscopic images at the point of no recapture of the device may cause us misjudge the accurate implantation depth owing to the parallax between the aortic annulus and delivery catheter. Furthermore, the MS is also unable to be displayed by fluoroscopy.

Thereupon, we introduced a transjugular ICE in November 2021 to bring the PPI rate as close to zero as possible. ICE enables real-time direct visualization of the MS during THV implantation, and thus the operator can adjust the landing point at the MS with a repositionable THV.^21^ Although some cases showing the feasibility of ICE were recently reported from Japan,^21^ no comparative study is available on the impact of ICE-guided TAVI on the reduction of subsequent conduction disturbances. Our study demonstrated that the combination of cusp-overlap technique and transjugular ICE provided a predictable THV bottom landing in the MS and an extremely low PPI rate (0.9%) without increasing adverse outcomes, such as THV dislodgement and paravalvular leakage. Of note, the novel implantation method using ICE also achieved a significant reduction of PPI rate in patients with baseline RBBB or short MS length, which are known factors that carry a high PPI risk.^8, 22^ Meanwhile, it should be recognized that high implantation may compromise the feasibility of coronary access or redo TAVI. However, in this regard, the direct visualization of the MS by ICE may also help avoid unnecessarily high implantation while ensuring a low PPI rate, because a longer MS would provide more freedom for a deeper THV implantation without necessarily affecting the conduction system. Indeed, in our study, ICE-guided TAVI provided a predictable ΔMSID with low variability (**Figure 3**).

Anecdotal reports have shown that a supra-annular prosthetic position may provide better valve hemodynamics. This may be attributed to the higher risk for valve thrombosis secondary to a constrained THV frame with deeper implantation and subsequent relative immobility of the THV leaflets.^23–25^ As expected, we found that the cusp-overlap technique provided a higher THV position, resulting in a larger EOA and lower mean gradient at 30 days, compared with the conventional implantation technique using the only the 3-cusps view. PPM, which is defined by the value of the EOA, was also less frequently detected in the cusp-overlap group.

### Limitations

First, although we used our prospective institutional database, implantation depth was retrospectively assessed from post-deployment aortic angiograms. Although the measurement was performed by two independent cardiologists blinded to clinical outcomes to mitigate bias, the implantation technique selected could be easily identified on the angiograms. Second, a bias based on the learning curve of THV implantation should also be considered because of the historical controlled nature of our study. Fortunately, baseline characteristics reported to be predictive for PPI, such as the MS length, prosthesis oversizing, and the prevalence of pre-existing RBBB and first-degree AVB, did not significantly differ between groups, whereas the procedure time and contrast volume were actually greater in the conventional “3-cusps view without ICE” group. Third, our data set only included information up to 30 days, and the impact of ICE-guided TAVI on longer-term clinical outcomes cannot yet be discussed. Fourth, MS length is an anatomic surrogate of the His bundle location, but the exact location of the His bundle may slightly differ, as has been reported.^26^ Finally, our examinations focused solely on the Evolut THV series. Theoretically, TAVI with other commercially available repositionable THV is expected to benefit from ICE guidance; however, further studies are necessary before applying this method to other THVs.

### Conclusions

A high risk for PPI remains a concern following self-expandable TAVI despite continued improvement in device and implantation methodology. Our novel method combining cusp-overlap view and ICE achieved a reliably higher THV position with substantial reduction in the subsequent rates of PPI and conduction disturbances without compromising procedural safety and THV hemodynamics. In particular, real-time direct visualization of the MS using ICE should be recognized as an important technique to improve patient outcomes.

### What Is Known?

A high permanent pacemaker implantation (PPI) risk remains a concern of self-expandable transcatheter aortic valve implantation (TAVI), despite continued improvement of implantation methodology, including cusp-overlap technique.

### What the Study Adds?

Transjugular intracardiac echocardiography (ICE) enables a real-time direct visualization of membranous septum. Our novel method combining cusp-overlap technique and ICE achieved a reliably higher transcatheter heart valve (THV) position with substantial reduction in the rates of PPI and conduction disturbances after TAVI without compromising procedural safety and THV hemodynamics.

## Data Availability

The data that support the findings of this study are available from the corresponding author, KI, upon reasonable request.

## Acknowledgments

None.

## Supplemental Material

Figures S1-S2

## Figure legends

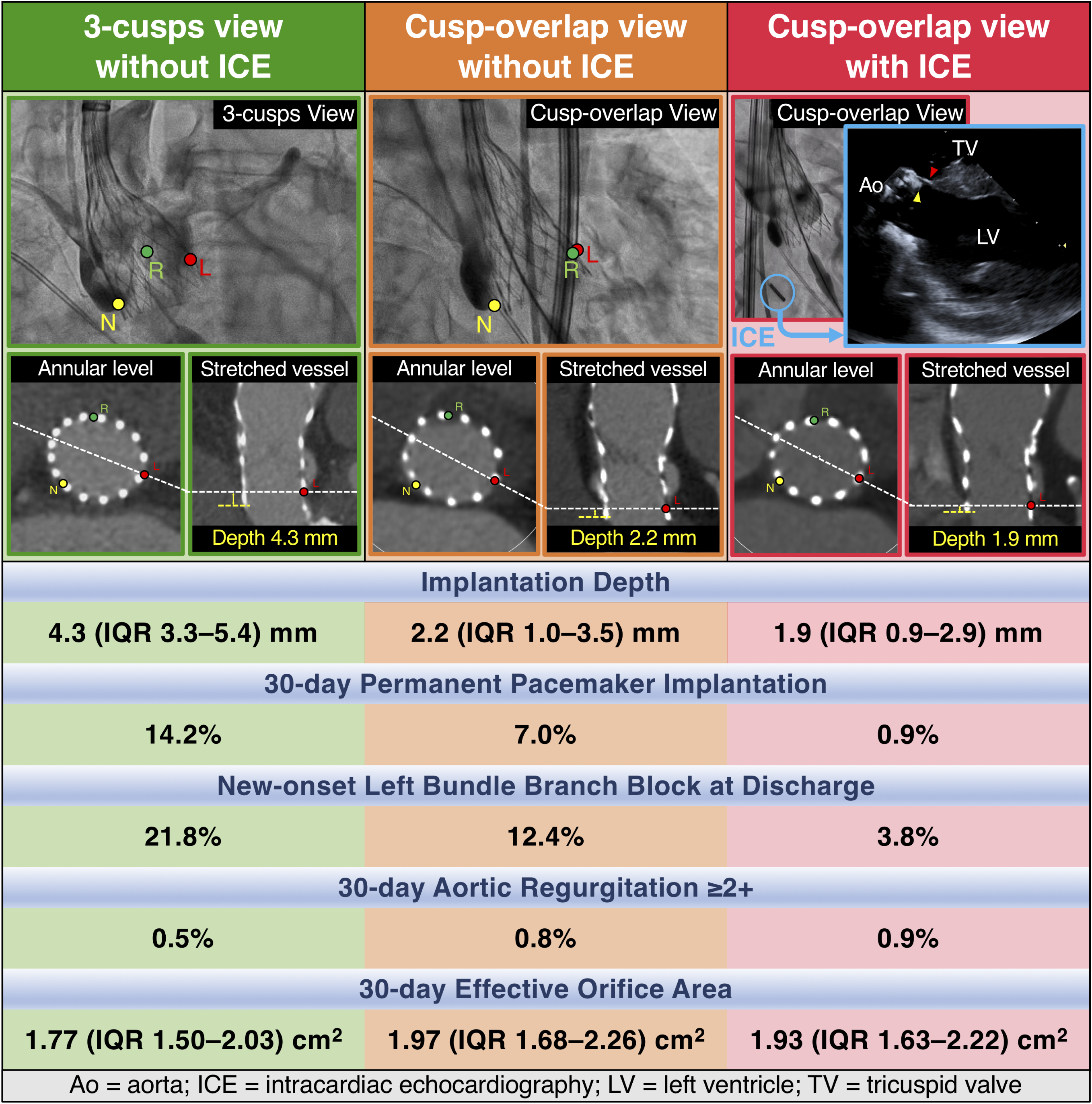

